# Physical Activity, Nutritional Status, and Cardiovascular Disease Prevalence in Geriatric Outpatients: A Cross-sectional Study in Swabi

**DOI:** 10.64898/2026.07.05.26355867

**Authors:** Saman Batool, Saddis Khan, Tanzeela, Abida Noreen, Usama Fahad, Azhar Ayub

## Abstract

**Introduction:** Cardiovascular diseases (CVD) are the leading cause of death among older adults worldwide. Cardiovascular risk is high in older adults, with modifiable risk factors including physical inactivity and poor diet.

**Objectives:** To evaluate geriatric physical activity, nutritional status (Mini Nutritional Assessment (MNA)), and CVD prevalence, and to explore associations between physical activity and nutrition with cardiovascular risk.

**Methodology:** A cross-sectional study included 150 geriatric participants. A structured questionnaire used a WHO-aligned physical activity assessment tool (GPAQ) and the validated six-item MNA screening tool. Statistical analysis was performed using IBM SPSS 22.0.

**Results:** Among participants, 56.7% were physically inactive and 43.3% were active. CVD was present in 50.7% of participants. The mean MNA score was 8.83 ±1.98, indicating that a substantial proportion of participants were at risk of malnutrition. Physically active individuals had significantly lower odds of CVD compared with inactive participants (OR = 0.42, 95% CI: 0.21-0.84, p = 0.014). Nutritional risk was also significantly associated with increased prevalence of cardiovascular disease (p < 0.05).

**Conclusion:** Physical inactivity and nutritional vulnerability are highly prevalent in this geriatric cohort and are significantly associated with CVD. The association was 58% lower odds of CVD (OR=0.42, 95% CI: 0.21–0.84), though the wide interval precludes a precise estimate of effect magnitude.

## Introduction

Cardiovascular diseases (CVDs) remain the foremost cause of global morbidity and mortality, with an estimated 19.2 million deaths in 2023 and over 437 million disability-adjusted life-years (DALYs). The global prevalence of CVD has more than doubled since 1990, rising from 311 million to 626 million cases in 2023, driven by population aging, urbanization, and persistent exposure to modifiable cardiometabolic risk factors. (1)

Pakistan, with its huge population of the 5th most populous country in the world, faces an alarming cardiovascular epidemic. The number of CVD cases increased by 110% during 1990-2019 (from 4.1 to 8.6 million cases), and the number of DALYs due to CVD increased by 119% during the same period. (2) A cross-sectional study in 53 cities in Punjab reported an overall 17.5% prevalence of CVDs in the general population of the city. (3) A number that is significantly higher in the elderly because of lifetime exposure to multiple cardiovascular risk factors. According to the World Health Organization Eastern Mediterranean Region (WHO EMRO) 2022 report, hypertension and diabetes are the top two metabolic risk factors for CVD morbidity in Pakistan. (4)

Of these, two behavioral and nutritional factors have been the focus of growing scientific interest as potentially modifiable risk factors for geriatric CVD: physical activity level and nutritional status. Physical inactivity is the fourth biggest risk factor for death around the world, causing 3.2 million deaths per year, according to the World Health Organization. (5) At the same time, 20-50% of community-dwelling and hospitalized older people worldwide are affected by malnutrition, defined as protein-energy undernutrition, micronutrient deficiencies, and involuntary weight loss, due to musculoskeletal deterioration, reduced cardiorespiratory reserve, fear of falling, and social withdrawal. (6)

The best-validated and shortest geriatric-specific nutritional screening tool is the Mini Nutritional Assessment (MNA), with a sensitivity of 89% and a specificity of 81.8%.(7) The MNA has geriatric-specific assessment questions for nutritional and health status, independence, quality of life, cognition, mobility, and subjective health.(8) Despite this, the MNA is still not routinely used in the care of the elderly in Pakistan and is a diagnostic opportunity as well as an opportunity for early nutritional intervention.

Physical activity and nutritional status do not exist in isolation but rather are a tripartite, mutually reinforcing entity with cardiovascular risk. The protective effect of exercise on CVD risk factors includes improved endothelial function, anti-inflammatory effects, improved lipid profiles, lower blood pressure, and improved insulin sensitivity. (9)

When both are deficient, as is likely to be the case in a significant number of elderly Pakistanis, the associated cardiovascular risk may be multiplicative instead of additive, thus hindering each other in their effects on cardiovascular pathogenesis.(10) If both are deficient, as is likely to be the situation in a significant number of elderly Pakistanis, the cardiovascular risk may be multiplicative instead of additive, interacting in such a way as to hinder each other in their effects on cardiovascular pathogenesis.

Although there is an increasing evidence base from around the world, there is limited data available on the co-occurrence and interaction of physical inactivity, malnutrition, and cardiovascular disease (CVD) in geriatric populations in Pakistan. This study attempts to fill this gap by performing an integrated cross-sectional evaluation of physical activity, nutritional status using MNA, and prevalence of CVD and risk factor profile.

## Methodology

The study was conducted in the District Headquarters (DHQ) Hospital, Swabi, Khyber Pakhtunkhwa (KPK), Pakistan, which is a large government tertiary health care center and the main referral institution for this district. Ethical approval was obtained from the Ethical Review Board of Women University Swabi and the Research Ethics Committee of DHQ Hospital Swabi. The study was conducted in accordance with the Declaration of Helsinki (2013). Written informed consent was obtained from all participants.

### Participants

Consecutive, non-probability sampling was used until the target sample of 150 was reached. The sample size was calculated using the formula.

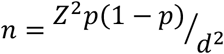

Based on an estimated CVD prevalence of 50% in South Asian geriatric populations, 95% confidence level, and 8% margin of error.

### Inclusion criteria

age >=60 years; residency in the study area >=1 year; capacity to provide informed consent; willingness to participate.

### Exclusion criteria

refusal to consent; severe cognitive impairment; acute illness at recruitment; physical disability precluding anthropometric measurement; inability to communicate in Urdu or the local language.

### Data collection

A structured interviewer-administered questionnaire was developed based on the Global Physical Activity Questionnaire (GPAQ) and was administered in Urdu and English and validated as being conceptually equivalent by bilingual experts.(11) The questionnaire consisted of four different sections: socio-demographics, medical history and the presence of cardiovascular diseases, physical activity, MNA-SF, dietary habits, and cardiovascular risk factors.

### Physical activity classification

Physical activity was assessed using self-reported frequency (days/week), session duration, type, and intensity. Participants reporting physical activity on ≥3 days/week were classified as Physically Active; those with <3 days/week as Physically Inactive, consistent with WHO Global Recommendations on Physical Activity for Health (2020) for older adults. (5)

### Nutritional status (MNA-SF)

The validated six-item MNA Short Form was administered to all participants. Items assessed: (1) decline in food intake over 3 months (0-2); (2) weight loss over 3 months (0-3); (3) mobility (0-2); psychological stress or acute illness (0-2); neuropsychological problems (0-2); (6) BMI (0-3). Total scores range from 0-14: >=12 = normal; 8-11 = at risk of malnutrition; <=7 = malnourished. (12)

### CVD assessment

Participant self-report of past diagnosis by a physician was used to determine CVD status. Medical records, prescription registries, or diagnostic investigations were not used to provide independent verification. Participants reported presence, type (ischemic heart disease, stroke, heart failure, or other), duration, and current medications.

### Statistical analysis

Data were entered, cleaned, and analyzed using IBM SPSS Statistics Version 22.0 (IBM Corp., Armonk, NY, USA). Categorical variables are reported as frequencies and percentages; continuous variables as mean +/- SD. Associations between physical activity level and CVD, and between MNA nutritional category and CVD, were evaluated using Pearson Chi-Square tests with Odds Ratios (OR) and 95% confidence intervals. An independent-samples t-test was used to compare MNA scores between the CVD-positive and CVD-negative groups. A p-value <0.05 (two-tailed) was considered statistically significant.

## Results

The total number of geriatric individuals involved was 150, of which 52.0% were females and 48.0% were males. The average age was 72.1 years. Most were married (76.0%), had higher education (58.0%), and were employed (64.0%). Key clinical features are shown in Figure 1. There were 47.3% with hypertension, 42.7% with diabetes mellitus, and 27.3% with a family history of CVD. Most (64.0%) were currently on medications.

**Figure 1.**
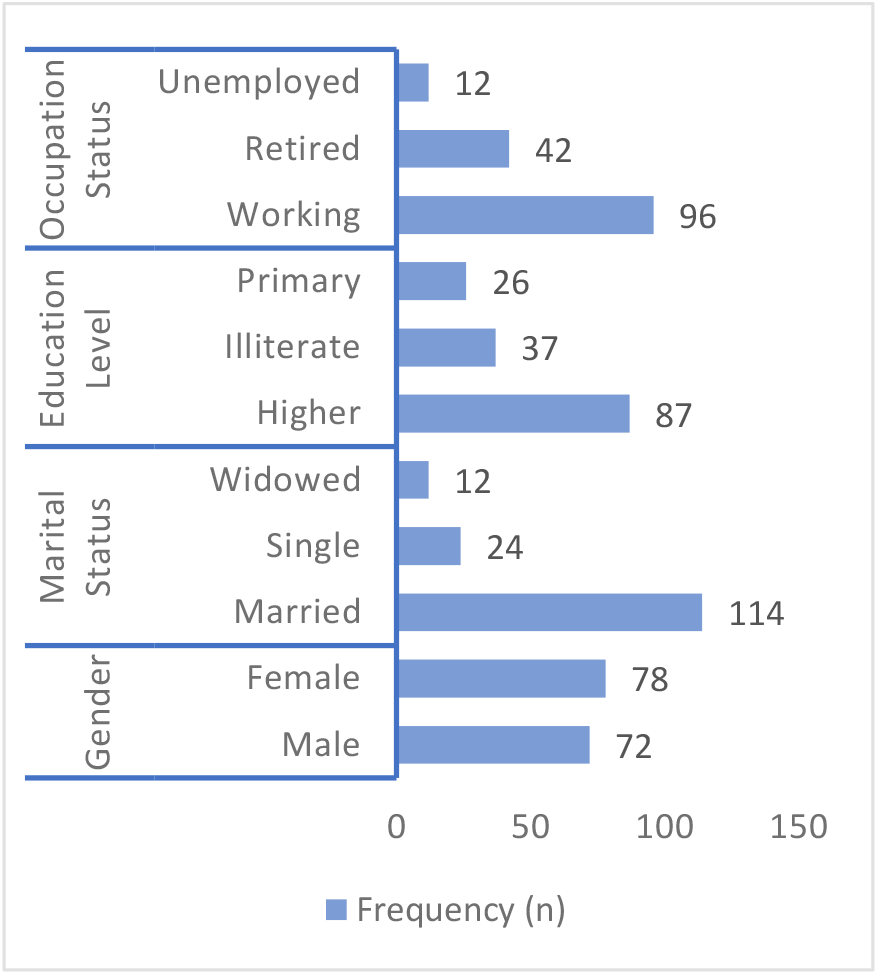
Frequency Distribution of Demographic Variables (N=150)

56.7% of the participants were physically inactive (Figure 2). Walking (45.3%) and household activities (40.0%) dominated, with very little formal exercise or participation in sport (4.0%). Almost one-fifth (19.3%) were completely sedentary. Most of those who were active participated at moderate intensity. Only 18.7% of the participants-maintained sessions that lasted more than 60 minutes.

**Figure 2.**
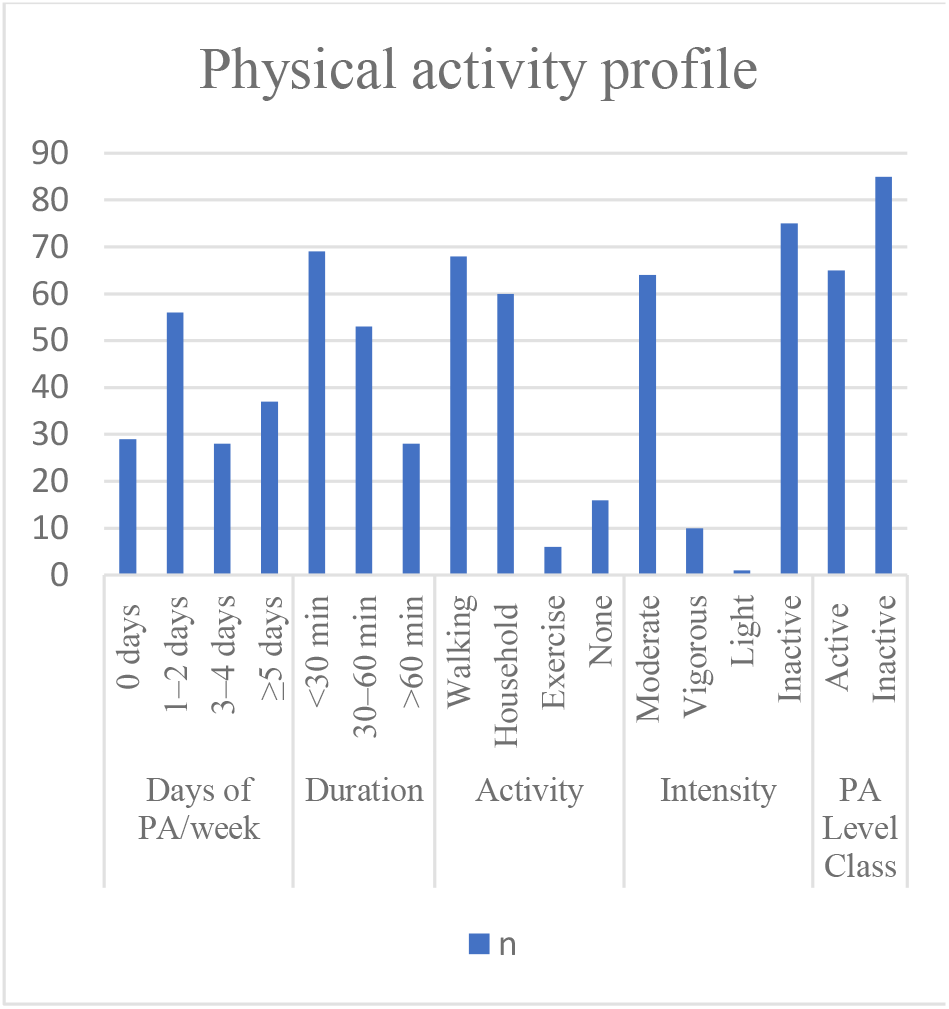
Physical Activity Profile of the Study Population (N=150)

The mean MNA score was 8.83 +/- 1.98, and the population-level average was in the ‘at risk’ category (Figure 3). Only 4.7% had normal nutritional status; 95.3% were either at risk of malnutrition (74.0%) or overtly malnourished (21.3%). The 25th percentile was 8.0, and the 75th percentile was 10.0, indicating that most participants clustered in the mid-to-lower nutritional risk range. Key contributors to nutritional risk included moderate-to-severe appetite decline (47.3%), recent weight loss (41.4%), psychological stress or acute illness (38.0%), and neuropsychological problems (33.4%).

**Figure 3.**
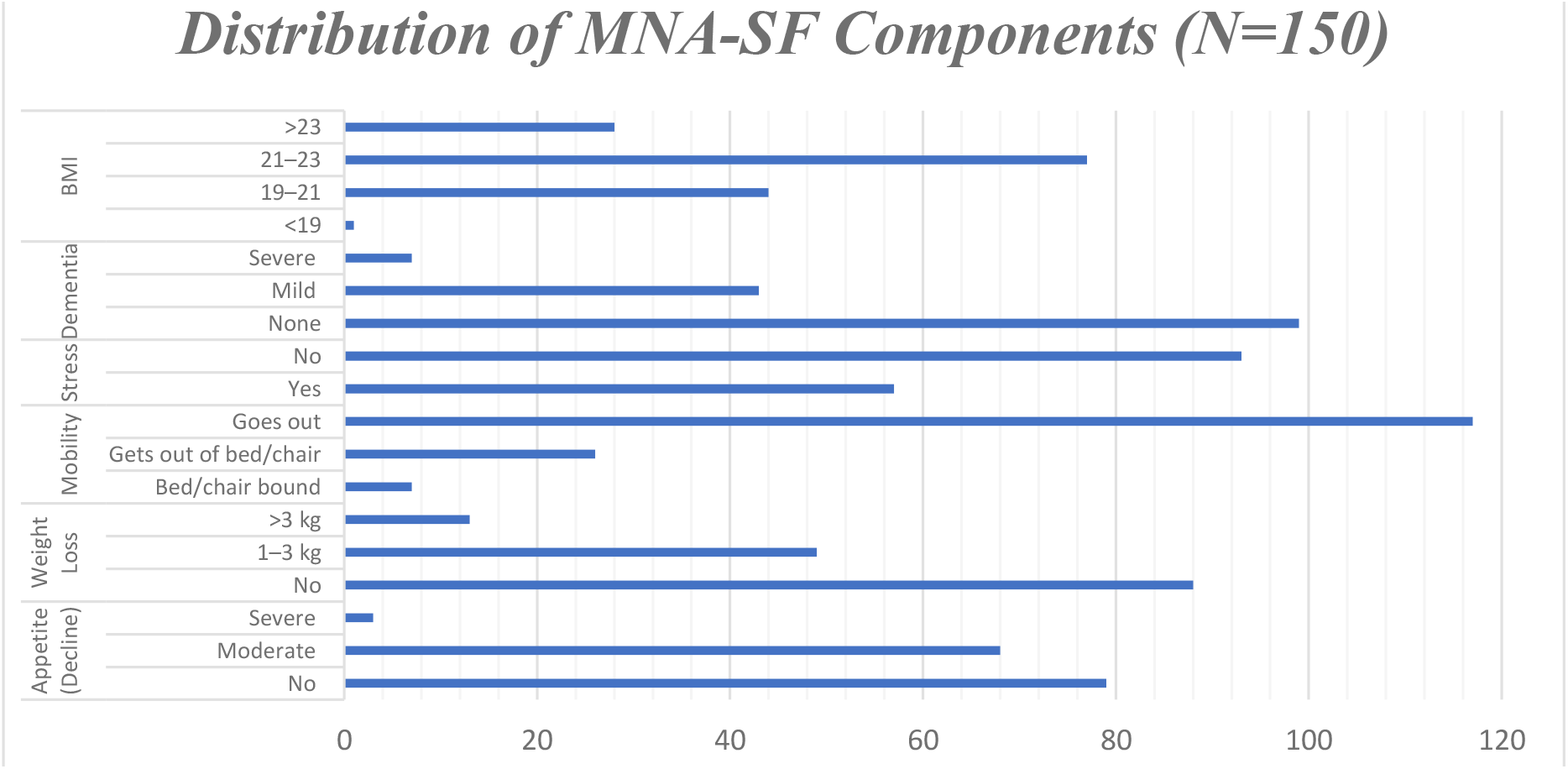
Distribution of MNA-SF Components (N=150)

CVD was present in 50.7% (n=76) of participants (Table 1). Stroke was the predominant subtype, accounting for 40.8% of CVD cases, followed by heart failure (25.0%) and ischemic heart disease (17.1%). High blood pressure was present in 47.3% of the total sample; elevated cholesterol in 34.0%. Smoking was uncommon; 90.0% had never smoked.

**Table 1.**
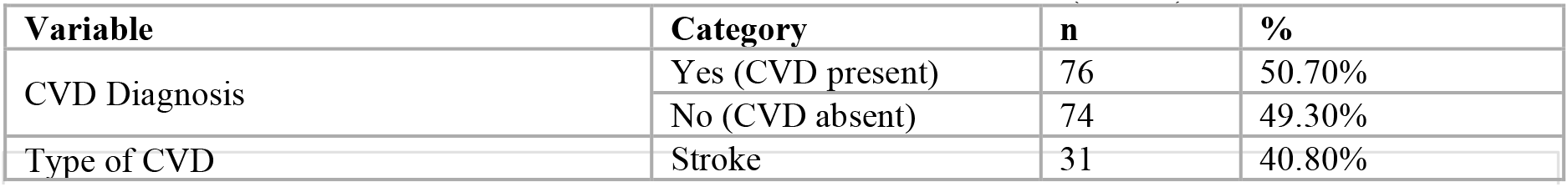
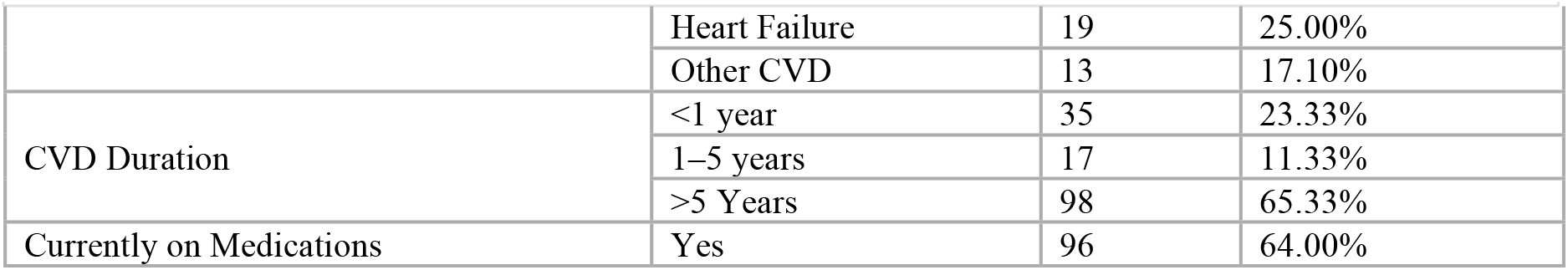
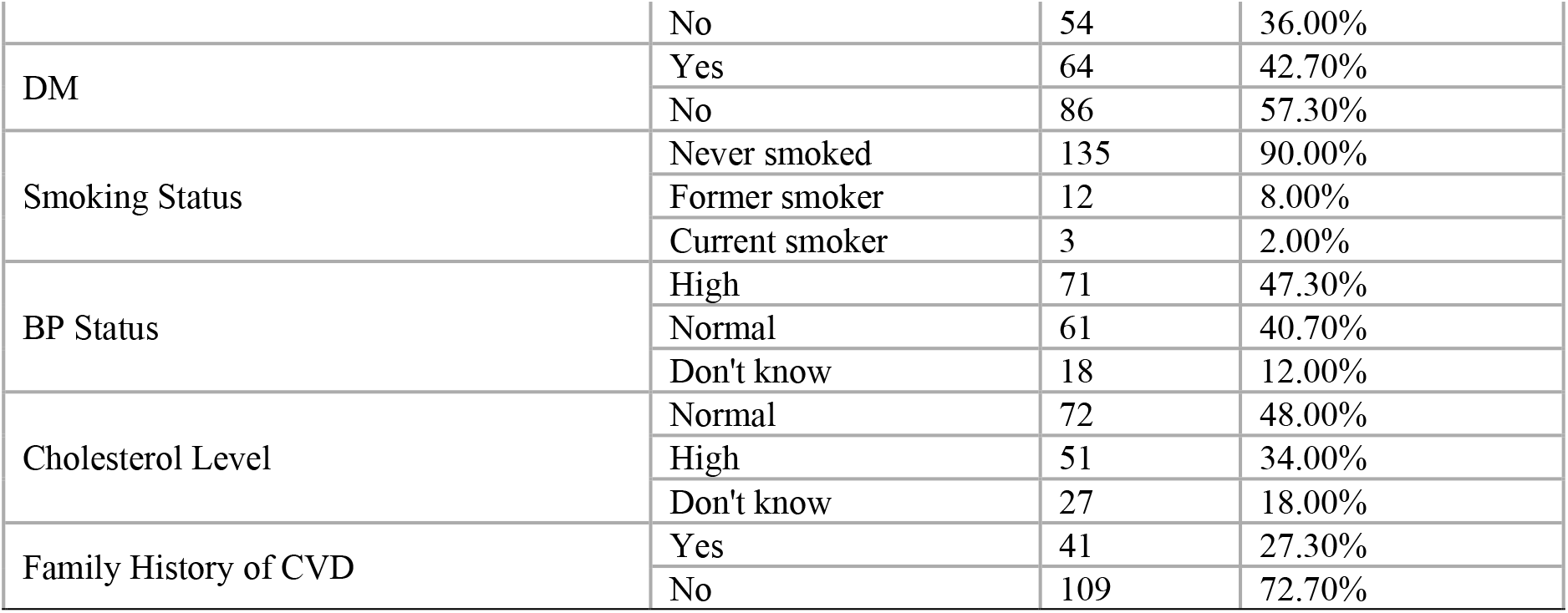
CVD Prevalence and Clinical Profile (N=150)

Physically active participants had a CVD rate of 38.5%, compared to 60.0% among inactive participants (p = 0.014; Table 2). The Odds Ratio of 0.42 indicates that physically active elderly individuals had 58% lower odds of CVD compared to their inactive counterparts. The association between PA frequency (days/week) and CVD was even stronger (p<0.001), consistent with a dose-response relationship.

**Table 2.** Bivariate associations of physical activity, nutritional status, and risk factors with CVD diagnosis (N=150)

| Risk Factor | CVD Yes n(%) | CVD No n(%) | Statistic/p-value |
| --- | --- | --- | --- |
| PA Active ($\geq 3$ days/wk) | 25 (38.5) | 40 (61.5) | $p = 0.014^*$ |
| PA Inactive ($< 3$ days/wk) | 51 (60.0) | 34 (40.0) | OR=0.42 |
| PA frequency (days/wk) | -- | -- | $p < 0.001^*$ |
| MNA Normal ($\geq 12$) | 3 (42.9) | 4 (57.1) | $p = 0.159$ |
| MNA At risk (8-11) | 52 (46.8) | 59 (53.2) |  |
| MNA Malnourished ($\leq 7$) | 21 (65.6) | 11 (34.4) | |
| Mean MNA score (t-test) | 8.47 ± 1.93 | 9.19 ± 1.98 | $t = -2.244$, $p = 0.026^*$ |
| Diabetes mellitus | 64(42.7) | 86(57.3) | $p < 0.001^*$ |
| Family history of CVD | 41(27.3) | 109(72.7) | $p < 0.001^*$ |
| Blood pressure (high) | 71(47.3) | 61(40.7) | $p < 0.001^*$ |
| Smoking status | 135(90) | 15(10) | $p = 0.185$ |
| Gender | -- | -- | $p = 0.749$ |
| Fatty food consumption | -- | -- | $p = 0.327$ |

\* $p < 0.05$ statistically significant. PA = physical activity; MNA = Mini Nutritional Assessment; OR = Odds Ratio

When the MNA score was analyzed as a continuous variable, CVD-positive participants had significantly lower mean scores than CVD-negative participants (8.47 vs. 9.19; t=- 2.244, p=0.026).

The strongest clinical predictors of CVD diagnosis in this cohort were blood pressure status (p<0.001), family history of CVD (p<0.001), and diabetes mellitus (p<0.001). In this population, there was no significant association between gender, smoking status, or fatty food consumption and CVD.

While the three-category MNA classification did not reach statistical significance against CVD (p=0.159) likely attributable to the small Normal subgroup (n=7) a clinically notable gradient was evident: CVD prevalence was 42.9% in the Normal, 46.8% in the At-Risk, and 65.6% in the Malnourished category.

## DISCUSSION

This study offers comprehensive local data on the interaction between physical activity, nutrition, and CVD in the Pakistani geriatric population, which is grossly underrepresented in the cardiovascular literature. These three keys’ findings-high prevalence of physical inactivity and nutritional deprivation among older adults, high proportion of older adults with CVD, and physical activity and nutritional status being independently associated with CVD prevalence all point to the dire need for integrated geriatric cardiovascular prevention strategies in Pakistan.

The 56.7% physical inactivity rate is consistent with the epidemiological evidence from the world that shows how physical inactivity decreases significantly with aging. Milanovic et al. documented age-related decreases in physical activity due to musculoskeletal deterioration and reduced cardiorespiratory reserve. (13) The WHO estimates that over 31% of adults globally fail to meet minimum activity recommendations, a proportion that increases substantially after age 60. (5) In our cohort, the predominance of walking (45.3%) and household activities (40.0%) as the primary forms of exercise reflects both cultural norms and the limited availability of organized exercise facilities in this socioeconomic context, a pattern reported across South Asian and Middle Eastern geriatric populations.

The near-universal nutritional vulnerability in this cohort (95.3% at-risk or malnourished; mean MNA 8.83) exceeds typical rates of 20-50% reported in systematic reviews of community-dwelling elderly using the MNA, and likely reflects the combined burden of poverty, high comorbidity, limited dietetic services, and the multifactorial etiology of malnutrition in this group -- including appetite decline (47.3%), weight loss (41.4%), psychological stress (38.0%), and cognitive impairment (33.4%).

The absence of normal nutrition status (4.7%) in this sample is a notable finding and is a systemic issue, as the MNA is endorsed by more than 22 international expert bodies as a first-line geriatric screening tool. (14) and is yet almost nonexistent in routine geriatric care in Pakistan.

The prevalence of CVD in the geriatric hospital-based population was 50.7%. The association between the MNA and the risk of CVDs may be partially driven by MNA-SF (mobility), given the high prevalence of stroke (40.8%) and heart failure (25.0%) in the cohort. The diseases themselves may directly affect mobility, resulting in poor MNA scores that are not related to the nutritional status. This, however, may be a reflection of functional limitation due to disease, not nutrition, and this could account for the lower MNA scores of those with CVD (8.47 vs. 9.19, p = 0.026), as observed in other South Asian populations. (15) Diabetes was strongly associated with CVD (p<0.001), consistent with WHO EMRO data showing diabetes as a major metabolic risk factor for CVD morbidity in Pakistan. (4)

The main result that physically active geriatric individuals had 58% lower odds of CVD (OR=0.42, p=0.014, E-valve=) is clinically and policy-actionable. Certain highly prevalent conditions, such as diabetes mellitus and hypertension, were strongly associated with CVD in our study and may have led to an overestimation of the protective effect of physical activity by residual confounding on these conditions. The underlying mechanisms are well-established: physical activity reduces CVD risk through improved endothelial function, lower systemic inflammation, favorable lipid modification, blood pressure reduction, and enhanced insulin sensitivity.(9) These mechanisms are particularly important in geriatrics, where aging-related arterial stiffening and endothelial dysfunction already predispose individuals to cardiovascular pathogenesis. The finding is reinforced by Wu et al., who reported a Hazard Ratio of 0.70 for CVD in long-term physically active populations and demonstrated a significant inverse dose-response relationship between PA frequency and cardiovascular mortality in 77,541 older adults. (16) The graded association between PA frequency and CVD (p<0.001) seen in our data further supports setting incremental physical activity goals in geriatric cardiovascular prevention, even a shift from zero to one or two activity days per week may confer measurable risk reduction.

With 67.3% of the cases of CVD having a disease duration of less than 1 year, the 3-month recall period in the MNA likely had a period of overlap with early disease. This is a time of reduced intake and often catabolic stress and this can lead to a decrease in MNA scores. This is likely to be disease-related malnutrition because MNA scores were significantly lower in the CVD-positive participants (8.47 vs. 9.19, p = 0.026) compared to the malnourished participants. Because of the cross-sectional design, no causal inferences can be drawn from the observations, and the relationship between the two might be either one-way or two-way.(17,18) The 12-year prospective data from Li et al. Demonstrating that MNA-identified malnutrition significantly predicts CVD mortality after adjustment for conventional risk factors provides particularly strong prognostic validation for our cross-sectional finding.(19) The non-significant categorical MNA result (p=0.159) is most plausibly explained by the very small Normal nutritional status group (n=7), which substantially limits statistical power for the Chi-Square analysis; the biological gradient CVD prevalence: Normal 42.9%, At-Risk 46.8%, Malnourished 65.6% is nonetheless clinically compelling.

Limitations: The self-reported CVD diagnoses and physical activity data may introduce misclassification or recall bias. The cross-sectional study design does not allow for causal inferences, and the sample is drawn from a single-center hospital, which might not be generalizable. Measures should be clinically validated, and multicenter cohorts and studies should be performed in the future.

## CONCLUSION

Physical inactivity and nutritional vulnerability are nearly universal in this geriatric Pakistani cohort and are significantly associated with cardiovascular disease. Active elderly individuals had 58% lower odds of CVD compared to inactive peers, and lower MNA scores were significantly associated with CVD presence. Diabetes mellitus, hypertension, and family history remain the strongest clinical predictors. These findings provide locally generated evidence supporting the integration of routine MNA nutritional screening and structured physical activity promotion into geriatric primary care in Pakistan. A multidisciplinary, multidomain model of geriatric cardiovascular prevention encompassing physical activity promotion, nutritional support, and cardiometabolic risk management offers the most promising pathway toward reducing the CVD burden in this vulnerable population.

## Data Availability

All data produced in the present study are available upon reasonable request to the authors

## ACKNOWLEDGEMENTS

The authors thank the administration and staff of DHQ Hospital, Swabi, for facilitating data collection, and all geriatric participants for their voluntary participation.

